# Shop-to-Stop Hypertension Statistical Analysis Plan

**DOI:** 10.1101/2025.10.30.25339100

**Authors:** Joyce Deng, Anthony Devaux, Chris Gianacas, Aletta E. Schutte

## Abstract

**Background:** High blood pressure (BP) is the leading global risk factor for death. In Australia, BP control has stagnated, with approximately 50% awareness among those with high BP. Shop-to-Stop Hypertension evaluates whether retail-based screening plus digital nudges can improve detection and follow-up. This document extends the already-published study protocol by pre-specifying the planned analyses.

**Design and Setting:** Multi-centre, parallel-group, cluster-randomised clinical trial across 30 Bunnings stores in New South Wales, Australia. SiSU Health Stations will be installed in the participating Bunnings stores to screen the public, with stores randomised to whether participants at that store receive text message nudge to retest or not.

**Outcomes:** The primary outcome is proportion of patients having a repeat BP check at a SiSU Health Station in the text-message based nudge group versus usual care. Secondary outcomes also compare between these two groups and include examining change in BP control (continuous and binary), change in weight, body mass index, and body fat percentage (all continuous). Medication use, health service use, number of BP checks, and awareness of BP risks will all also be examined, as will factors associated with BP control and whether certain groups are more responsive to text message-based nudges.

**Planned analyses:** All analyses will be intention-to-treat with alpha=0.05 (two-sided), presented with 95% confidence intervals, and conducted using R software version 4.5. Categorical outcomes including primary will be presented as relative risks and analysed using robust Poisson mixed models with random intercepts for store and fixed effects include treatment arm, stratification variables (socioeconomic decile and rurality), and baseline SBP. Continuous outcomes will be analysed similarly using linear mixed models. In the case of convergence issues, cluster-level stratification fixed effects will be omitted with fallback to generalised estimating equation modelling. Multiplicity across six secondary efficacy endpoints will be controlled using Holm-Sidak and potential selection bias will be examined using tipping point analyses. Subgroup and sensitivity analyses include stratifying or controlling for key prognostic variables e.g., age, sex, site rurality, BMI, pregnancy, BP meds at baseline, and self-reported diabetes.

## 1 Study Background and analysis scope

The leading risk factor for death worldwide is raised blood pressure (BP)(1). In Australia alone, high BP accounted for 25,498 deaths (28% of all deaths) in 2019 (2). As such, better BP control could save more lives than any other single treatment intervention (3). However, BP control rates have stagnated in Australia since 2011 with only one in two people with high BP being aware of their condition based on the results of the 2017-2019 May Measurement Month BP screening campaign (4). BP control rates could be improved if a larger proportion of the population were screened and high BP detected for further care, leading to the initiation of effective treatment and potentially saving thousands of lives.

This study is a multi-centre, cluster randomized clinical trial which investigates the use of a novel approach to improve awareness and detection of high BP. The general public will be able to screen their BP and related health markers at SiSU Health Stations (SiSU Health Group, Australia). These SiSU Health Stations will be located within 30 Bunnings stores across New South Wales, Australia. Bunnings stores will be randomized in a 1:1 ratio to the intervention vs. control groups. Participants screened with high BP (>140 and/or >90 mmHg) at a store randomized to the intervention group will receive mobile text message-based nudges encouraging repeat BP checks and encouraging them to visit their general practitioner (GP) in addition to the usual protocol of the SiSU Health Station. The usual protocol includes a screen prompt to check their BP with their GP as well as sending the participant an email indicating a high BP reading and encouraging the participant to visit their GP for BP management. Participants screened at a store randomized to the control group will not receive the mobile text message-based nudges – only the usual protocol. Note that it is possible for individuals to perform measurements at a health station and at a point of care. Only the measurements taken at a health station are considered in this study.

The goal of this study is to improve awareness and detection of high BP in Australia. Current evidence shows simple text message-based nudges can be used as an unobtrusive, low-cost method to encourage health promotion (5).

## 2 Aims

The primary aim of the Shop-to-Stop Hypertension study is to determine whether text message-based nudges encouraging repeat BP checks at SiSU Health Stations and encouraging participants to visit their GP will result in a greater proportion of participants with detected high BP as measured at SiSU Health Stations rechecking their BP at a SiSU Health Station over the duration of the study compared to those who do not receive text message-based nudges.

Secondary aims include evaluating group differences in the change in systolic and diastolic BP, weight, body mass index (BMI), body fat percentage, use of BP medications, health risk behaviours, visits to the GP, and number of visits to SiSU Health Stations. In addition, this study aims to determine which factors are associated with improved BP control as well as whether certain groups of individuals are more responsive to text message-based nudges.

## 3 Hypotheses

We hypothesize that the addition of text message-based nudges to the usual protocol at SiSU Health Stations will be superior compared to the usual protocol in terms of promoting repeat health checks.

## 4 Outcomes

Primary:

1. Difference in proportion of participants with high BP detected having a repeat BP check at a SiSU Health Station in the text message-based nudges vs. usual care groups reported as a risk ratio (RR) with 95% confidence interval (CI) and p-value.

Secondary:

1. Efficacy:

a. Net difference in change in systolic BP (mmHg) from baseline to last repeat measurement on a SiSU station reported as a difference in means with 95% CI and p-value
b. Net difference in change in diastolic BP (mmHg) from baseline to last repeat measurement on a SiSU station reported as a difference in means with 95% CI and p-value
c. Difference in proportion of participants achieving BP target of <140/90 mmHg at final repeat measurement on a SiSU station reported as an RR with 95% CI and p-value
d. Net difference in change in weight (kg) from baseline to last repeat measurement on a SiSU station reported as a difference in means with 95% CI and p-value
e. Net difference in change in BMI from baseline to last repeat measurement on a SiSU station reported as a difference in means with 95% CI and p-value
f. Net difference in change in body fat percentage from baseline to last repeat measurement on a SiSU station reported as a difference in means with 95% CI and p-value
2. Medication Use:

a. Difference in proportion of participants with a self-reported change in BP medications reported as an RR with 95% CI and p-value
b. Difference in proportion of participants with a self-reported change in health risk behaviours at their final repeat measurement on a SiSU station reported as an RR with 95% CI and p-value
3. Health Service Use and Engagement:

a. Difference in proportion of participants who had self-reported medical attendance with their GP for high BP management reported as an RR with 95% CI and p-value
4. Number of BP checks, awareness of BP risks:

a. Difference in proportion of participants aware of high BP risks at final repeat SiSU measurement reported as an RR with 95% CI and p-value
b. Number of visits to a SiSU health station reported as a difference in means with 95% CI and p-value
5. Factors associated with improved BP control
6. Whether certain groups are more responsive to text message-based nudges

## 5 Other variables of interest

Participants are uniquely identified by a user ID in the system. After creating a user profile, subsequent measurements can be linked to their user ID. Measurements at SiSU health stations will be identified by the variable for the check type. Additionally, each individual SiSU station is identified by a unique station name.

At each SiSU measurement, information on a participant’s health risk behaviours is collected including the participant’s smoking habits (smoking on a daily basis, ever smoked more than 100 cigarettes), vaping habits (vaping on a daily basis), nutrition habits (daily vegetable or fruit intake), and physical activity (weekly physical activity). A change in health risk behaviours from baseline to the final repeat measurement at a SiSU station is defined as a change in response for any of the listed behaviours.

Each participant’s awareness of high BP risks is also collected at each measurement. Each participant is asked to select all the health conditions they are aware of which can be caused by high BP. Their awareness of high BP risks will be defined as the number of conditions they correctly select at each measurement, with incorrect selections reducing the count by one.

The time since a participant’s last visit to their GP will be collected and used to determine their self-reported medical attendance with their GP for high BP management. It is assumed a GP visit after the date of the participant’s first SiSU health measurement will have been for high BP management. The actual analysis endpoint will be any memory of a GP visit vs. no memory of a GP visit i.e., no attempt will be made to validate the amount of time in the past selected by the participant. Similarly, the participant’s response to whether they are currently taking medication for high blood pressure will be used to determine their self-reported change in BP medications. A change is defined as any change in response between the first SiSU measurement and final repeat SiSU measurement.

## 6 Sample Size

Based on current SISU use, we anticipate an average of 15 checks per station per operating day (conservative estimate). We target an average of 2.5 checks per participant per annum, equating to 163,350 total checks and 65,340 total participants. Assuming that 18% of those screened will have high BP (based on previous pilot data of two SiSU stations in Bunnings stores), this will equate to 11,700 people detected with raised BP.

A parallel two-group cluster-randomised design will be used to test whether the Group 1 (text message intervention) proportion (P1) is different from the Group 2 (control) proportion (P2) (H0: P1 - P2 = 0 versus H1: P1 - P2 ≠ 0). The comparison will be made using a two-sided Z-Test (Unpooled) based on the proportion difference, with a Type I error rate (α) of 0.05. The control group proportion (P2) is assumed to be 0.2. Recruiting 11,700 participants and assuming 70% drop out, results in 3,510 randomised participants completing follow-up. For a power of 90% with 15 clusters of 117 participants per cluster in Group 1 and 15 clusters of 117 subjects per cluster in Group 2, the detectable proportion difference (P1 - P2) is 0.085 (or P1 of 0.285). The intra-cluster correlation coefficient assumed is 0.02. That is, this trial will achieve 90% power to detect an 8.5% difference in the proportion of participants returning for a SiSU health station check. The detectable proportion difference (P1 - P2) was computed using PASS 2023, version 23.0.1.

## 7 Analyses

All analyses will be conducted on an intention-to-treat basis. All tests will be two-sided at a 0.05 level of significance (α = 0.05). All analyses will be conducted using R software version 4.5.

### 7.1 Multiplicity

The primary efficacy outcome will be analysed without multiplicity adjustment. Whether or not the primary outcome is significant, testing will proceed to the secondary efficacy outcomes. As the six secondary efficacy outcomes consider alternate characterisations of the primary efficacy outcome (reduction in BP) or a different efficacy measure (body mass/composition), control of the family-wise error rate is required. For the three secondary clinical outcomes, we will control the family-wise error rate by applying a sequential Holm-Sidak correction (6).This consists of ordering the derived p-values from smallest (most significant) to largest (least significant), and then comparing each to an adjusted level of significance calculated as 1-(1-0.05)1/C, where C indicates the number of comparisons that remain. The procedure halts as soon as a p-value fails to reach the corrected significance level.

The remaining endpoints are exploratory in nature and will not be adjusted for multiple comparisons.

### 7.2 Main analysis

Baseline characteristics will be presented by treatment group. Discrete variables will be summarized by frequencies and percentages. Continuous variables will be summarized using the means with the standard deviations and/or the medians with the interquartile ranges.

A robust Poisson mixed model with random intercepts for the store will be used to analyse the primary outcome, subsequent measurement at a SiSU station (yes/no). The model will include fixed effects for the treatment group as well as variables used to stratify Bunnings stores (i.e. IRSAD decile and rurality) and baseline SBP. The treatment effect will be presented using RRs with corresponding 95% CIs and p-values. Additionally, the ICC with its corresponding 95% CI will be reported.

Due to the nature of the primary outcome, missing data is not expected – participants will either have a subsequent measurement at an SiSU station or no subsequent measurements. Consequently, all analyses will be complete case.

In case of convergence issues using mixed model regression, cluster-level stratification fixed effects will be omitted from the mixed model in the first instance. Ongoing convergence issues will be addressed by using Generalized Estimating Equations (GEE) clustered by store instead. If convergence issues remain unresolved, a Gaussian GEE model will be considered (7).

### 7.3 Secondary efficacy analyses

While of clinical importance, the secondary efficacy endpoints present a risk of selection bias because they are only collected when a participant presents for one or more repeat BP checks, which is precisely the primary endpoint the intervention aims to influence. As such, in the first instance all secondary endpoints will be analysed as-is using the sequential Holm-Sidak correction discussed above noting this risk of bias.

To quantify the potential impact of differential missingness, a tipping point analysis will be conducted for each of the six secondary efficacy outcomes. This will consist of using multiple imputation (MI) to impute a secondary outcome at one repeat BP check for those patients who have no repeat BP checks recorded (across both treatment arms), with the addition of a shift parameter that adjusts the imputed values independently in each treatment arm. The secondary efficacy endpoint analyses are then rerun on the multiply imputed dataset, results are recorded, and the process runs again. By systematically varying the deltas associated with the control and intervention arms it is possible to produce a matrix of shift parameters and the resulting p-values. These matrices will then be presented graphically. For comparability, the tipping point analyses will be presented with a reference alpha of 0.05 i.e., not using the sequential Holm-Sidak correction.

### 7.4 Subgroup and Sensitivity Analyses

Additional analyses will be performed by adding the following covariates to the robust Poisson mixed model for the main analysis as fixed effects: age, sex, postcode-based rurality (rural-remote vs metropolitan based Bunnings), BMI, pregnant (if female), already taking BP meds at baseline, and self-reported diabetes. If important imbalances are noted in the baseline characteristics, the unbalanced variables will also be included as fixed effects in a second adjusted model along with the variables included in the first adjusted model listed above. The adjusted treatment effect will be presented as adjusted RRs with corresponding 95% CIs.

The following subgroups will be analysed: Males vs females; rural-remote vs metropolitan; age group; IRSAD; pregnant vs non-pregnant (females); body mass index categories; blood pressure categories; self-reported high BP in the last 12 months vs not; history of last seeing a GP (6 months ago or more frequently vs 12 months or longer); antihypertensive users vs non-users. For each subgroup, summary measures, including the raw counts and percentage within each treatment arm, will be provided. To determine the treatment effect, the subgroup and its interaction with the treatment group will be added as fixed effects to the robust Poisson mixed model used for the main analysis. The treatment effect will be presented as RRs with corresponding 95% CIs.

### 7.5 Other analyses

The net difference in the systolic BP will be analysed using a linear mixed model with the results presented as the difference in the mean with the 95% CI and p-value. The model will include fixed effects for the treatment group, stratification variables (i.e. IRSAD decile and rurality), and the baseline measurement. Otherwise, the analytical approach will be the same as presented above for the primary outcome. The net difference in other continuous outcomes diastolic BP, weight, BMI, and body fat percentage will be analysed using the same modelling approach.

The difference in proportion of participants achieving the target BP, with a self-reported change in BP medications, with a self-reported change in health risk behaviours, and who had self-reported medical attendance will be analysed using the same modelling approach as the primary outcome.

## Data Availability

Data for the planned analyses described in this SAP are in the process of being acquired and the final data availability statement will be included with the publication of results. That said, we anticipate that all data produced in the current study will be made available upon reasonable request to the authors.

## Acknowledgements

Thanks to Prof. Laurent Billot for his invaluable methodological input on optimal analyses of cluster-randomised studies.

## List of Abbreviations

Abbreviation: Definition
BMI: Body Mass Index
BP: Blood Pressure
CI: Confidence Interval
GP: General Practitioner
RR: Risk Ratio

## 10 Mock Tables

**10.1 Figure 1.**
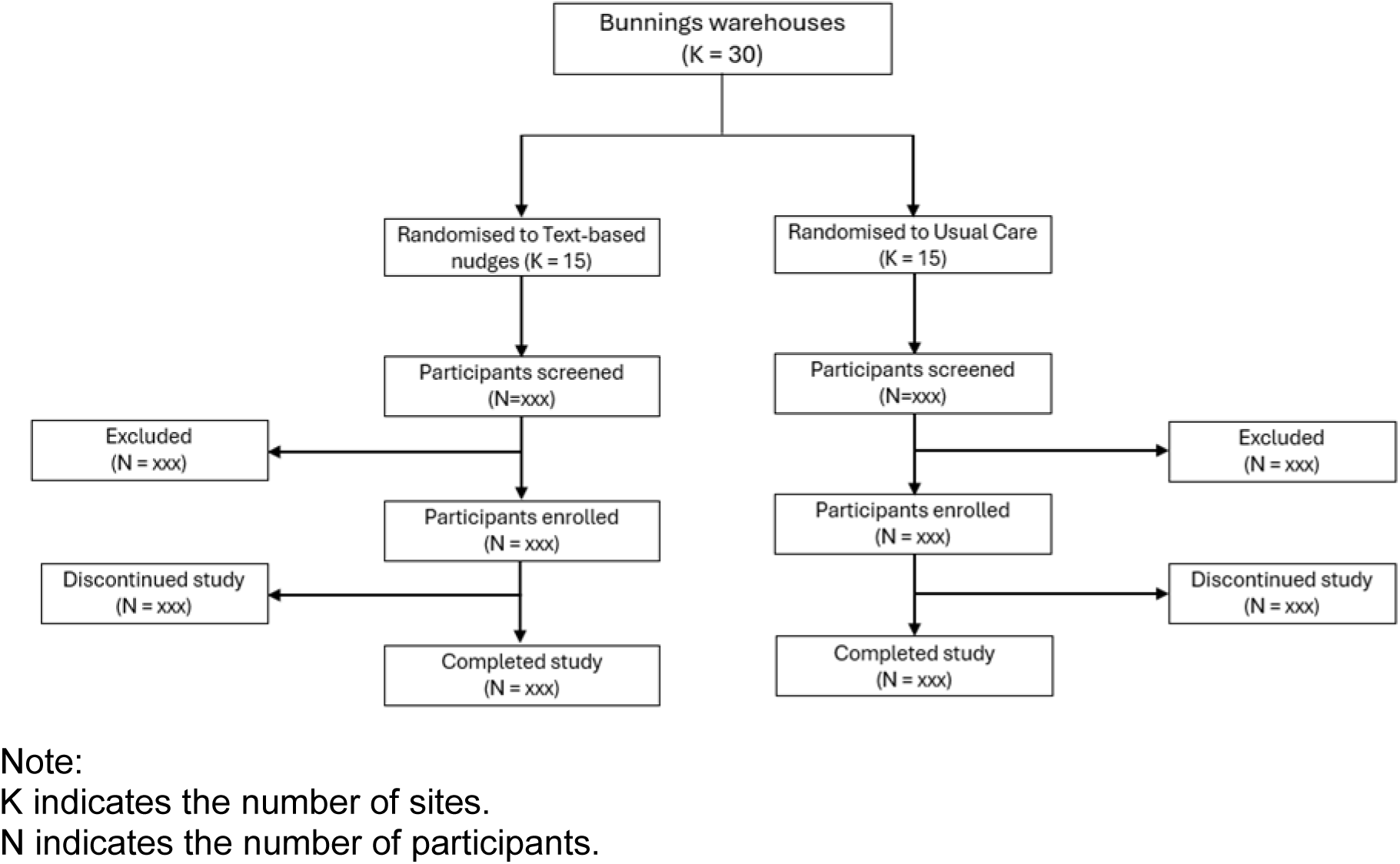
Consort Diagram (illustration)

**10.2 Table 1.**
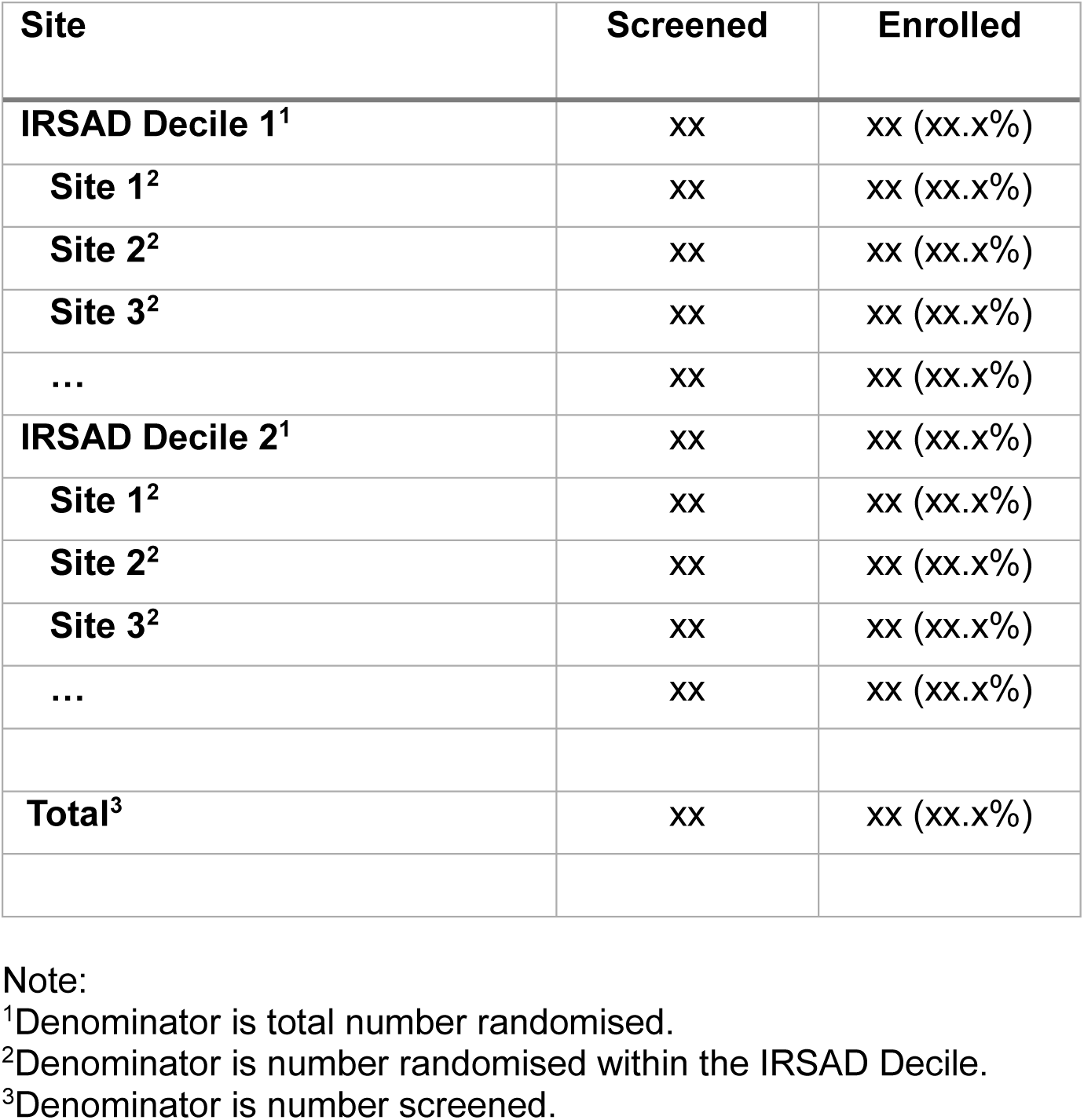
Enrolment by site

| Site | Screened | Enrolled |
| --- | --- | --- |
| **IRSAD Decile 1<sup>1</sup>** | xx | xx (xx.x%) |
| **Site 1<sup>2</sup>** | xx | xx (xx.x%) |
| **Site 2<sup>2</sup>** | xx | xx (xx.x%) |
| **Site 3<sup>2</sup>** | xx | xx (xx.x%) |
| ... | xx | xx (xx.x%) |
| **IRSAD Decile 2<sup>1</sup>** | xx | xx (xx.x%) |
| **Site 1<sup>2</sup>** | xx | xx (xx.x%) |
| **Site 2<sup>2</sup>** | xx | xx (xx.x%) |
| **Site 3<sup>2</sup>** | xx | xx (xx.x%) |
| ... | xx | xx (xx.x%) |
| **Total<sup>3</sup>** | xx | xx (xx.x%) |

Note:
<sup>1</sup>Denominator is total number randomised.
<sup>2</sup>Denominator is number randomised within the IRSAD Decile.
<sup>3</sup>Denominator is number screened.

**10.3 Table 2.**
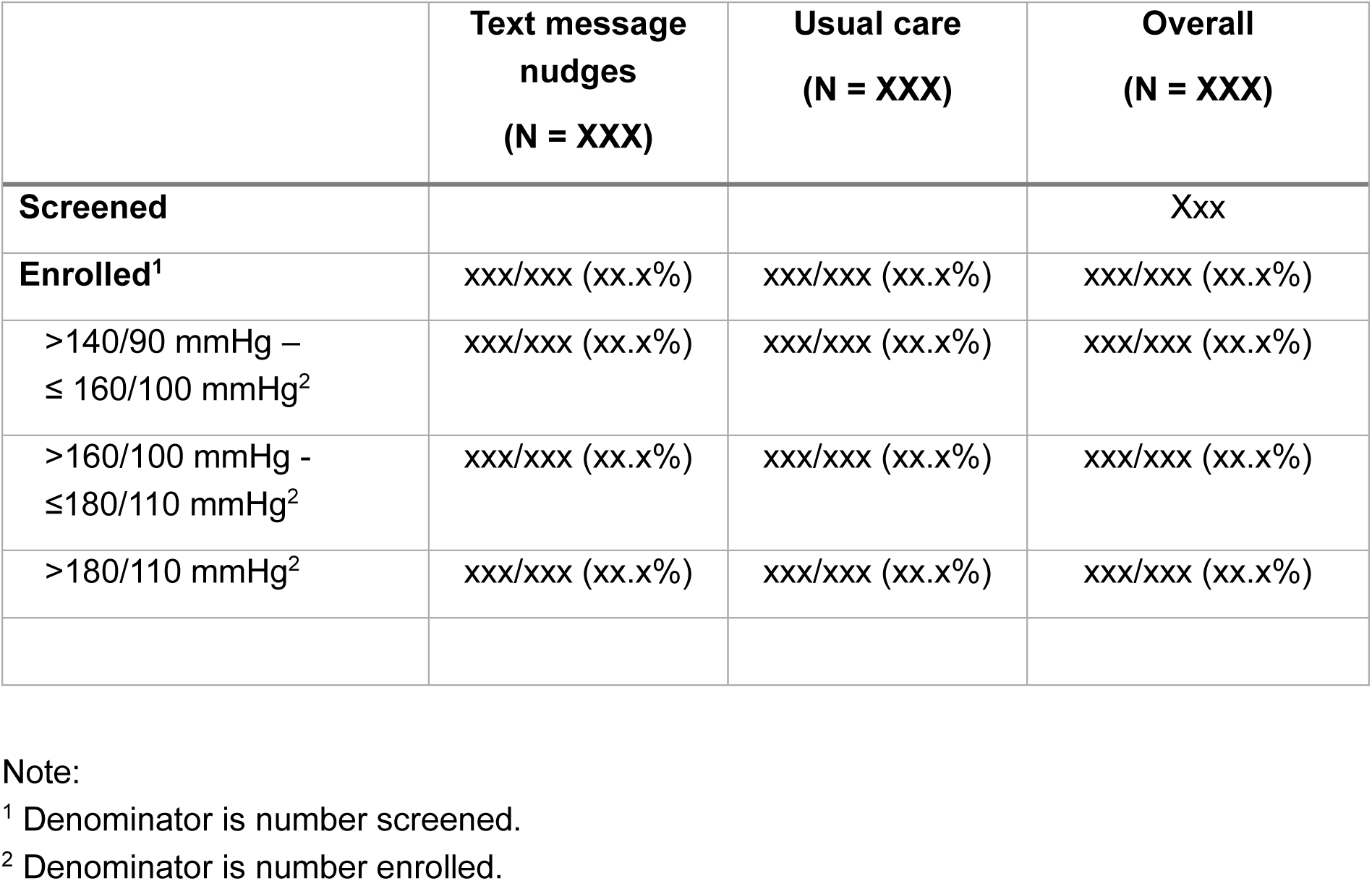
Patient disposition

|  | **Text message nudges**<br>**(N = XXX)** | **Usual care**<br>**(N = XXX)** | **Overall**<br>**(N = XXX)** |
| --- | --- | --- | --- |
| **Screened** |  |  | Xxx |
| **Enrolled<sup>1</sup>** | xxx/xxx (xx.x%) | xxx/xxx (xx.x%) | xxx/xxx (xx.x%) |
| >140/90 mmHg –<br>≤ 160/100 mmHg<sup>2</sup> | xxx/xxx (xx.x%) | xxx/xxx (xx.x%) | xxx/xxx (xx.x%) |
| >160/100 mmHg -<br>≤180/110 mmHg<sup>2</sup> | xxx/xxx (xx.x%) | xxx/xxx (xx.x%) | xxx/xxx (xx.x%) |
| >180/110 mmHg<sup>2</sup> | xxx/xxx (xx.x%) | xxx/xxx (xx.x%) | xxx/xxx (xx.x%) |

Note:
<sup>1</sup> Denominator is number screened.
<sup>2</sup> Denominator is number enrolled.

**10.4 Table 3.** Baseline characteristics

|  | **Text message nudges**<br>**(N=XXX)** | **Usual care**<br>**(N=XXX)** | **Overall**<br>**(N=XXX)** |
| --- | --- | --- | --- |
| **Sex at birth** |  |  |  |
| Male | xxx/xxx (xx.x%) | xxx/xxx (xx.x%) | xxx/xxx (xx.x%) |
| Female | xxx/xxx (xx.x%) | xxx/xxx (xx.x%) | xxx/xxx (xx.x%) |
| **Gender** |  |  |  |
| Man or male | xxx/xxx (xx.x%) | xxx/xxx (xx.x%) | xxx/xxx (xx.x%) |
| Woman or female | xxx/xxx (xx.x%) | xxx/xxx (xx.x%) | xxx/xxx (xx.x%) |
| Non-binary | xxx/xxx (xx.x%) | xxx/xxx (xx.x%) | xxx/xxx (xx.x%) |
| Prefer not to say | xxx/xxx (xx.x%) | xxx/xxx (xx.x%) | xxx/xxx (xx.x%) |
| **Age (years)** |  |  |  |
| N | xxx | xxx | xxx |
| Mean (SD) | xx.x (xx.x) | xx.x (xx.x) | xx.x (xx.x) |
| Median (IQR) | xx.x (xx.x; xx.x) | xx.x (xx.x; xx.x) | xx.x (xx.x; xx.x) |
| < 20 | xxx/xxx (xx.x%) | xxx/xxx (xx.x%) | xxx/xxx (xx.x%) |
| 20 - 24 | xxx/xxx (xx.x%) | xxx/xxx (xx.x%) | xxx/xxx (xx.x%) |
| 25 - 29 | xxx/xxx (xx.x%) | xxx/xxx (xx.x%) | xxx/xxx (xx.x%) |
| 30 - 34 | xxx/xxx (xx.x%) | xxx/xxx (xx.x%) | xxx/xxx (xx.x%) |
| ... |  |  |  |
| **Descent** |  |  |  |
| Aboriginal and Torres Strait Islander | xxx/xxx (xx.x%) | xxx/xxx (xx.x%) | xxx/xxx (xx.x%) |
| Pacific Islander or Maori | xxx/xxx (xx.x%) | xxx/xxx (xx.x%) | xxx/xxx (xx.x%) |
| None of the above | xxx/xxx (xx.x%) | xxx/xxx (xx.x%) | xxx/xxx (xx.x%) |
| **Region of birth** |  |  |  |
| Australia | xxx/xxx (xx.x%) | xxx/xxx (xx.x%) | xxx/xxx (xx.x%) |
| Asia | xxx/xxx (xx.x%) | xxx/xxx (xx.x%) | xxx/xxx (xx.x%) |
| Middle East | xxx/xxx (xx.x%) | xxx/xxx (xx.x%) | xxx/xxx (xx.x%) |
| North Africa | xxx/xxx (xx.x%) | xxx/xxx (xx.x%) | xxx/xxx (xx.x%) |
| Southern Europe | xxx/xxx (xx.x%) | xxx/xxx (xx.x%) | xxx/xxx (xx.x%) |
| Other | xxx/xxx (xx.x%) | xxx/xxx (xx.x%) | xxx/xxx (xx.x%) |
| **Remoteness area** |  |  |  |
| ... | xxx/xxx (xx.x%) | xxx/xxx (xx.x%) | xxx/xxx (xx.x%) |
| **SEIFA IRSAD decile** |  |  |  |
| 1 | xxx/xxx (xx.x%) | xxx/xxx (xx.x%) | xxx/xxx (xx.x%) |
| 2 | xxx/xxx (xx.x%) | xxx/xxx (xx.x%) | xxx/xxx (xx.x%) |
| 3 | xxx/xxx (xx.x%) | xxx/xxx (xx.x%) | xxx/xxx (xx.x%) |
| ... |  |  |  |
| **SEIFA IRSD decile** |  |  |  |
| 1 | xxx/xxx (xx.x%) | xxx/xxx (xx.x%) | xxx/xxx (xx.x%) |
| 2 | xxx/xxx (xx.x%) | xxx/xxx (xx.x%) | xxx/xxx (xx.x%) |
| 3 | xxx/xxx (xx.x%) | xxx/xxx (xx.x%) | xxx/xxx (xx.x%) |
| ... |  |  |  |
| **Self-reported waist circumference band (cm)** |  |  |  |
| Less than 102 cm | xxx/xxx (xx.x%) | xxx/xxx (xx.x%) | xxx/xxx (xx.x%) |
| 102 – 110 cm | xxx/xxx (xx.x%) | xxx/xxx (xx.x%) | xxx/xxx (xx.x%) |
| More than 110 cm | xxx/xxx (xx.x%) | xxx/xxx (xx.x%) | xxx/xxx (xx.x%) |

Note:
Denominators are number of participants with available data.

**10.5 Table 4.** Lifestyle Factors

|  | **Text message nudges**<br>**(N=XXX)** | **Usual care**<br>**(N=XXX)** | **Overall**<br>**(N=XXX)** |
| --- | --- | --- | --- |
| **Smoked 100 or more cigarettes in their life** |  |  |  |
| Yes | xxx/xxx (xx.x%) | xxx/xxx (xx.x%) | xxx/xxx (xx.x%) |
| No | xxx/xxx (xx.x%) | xxx/xxx (xx.x%) | xxx/xxx (xx.x%) |
| **Smokes at least daily** |  |  |  |
| Yes | xxx/xxx (xx.x%) | xxx/xxx (xx.x%) | xxx/xxx (xx.x%) |
| No | xxx/xxx (xx.x%) | xxx/xxx (xx.x%) | xxx/xxx (xx.x%) |
| **Smoking frequency** |  |  |  |
| Several times a day | xxx/xxx (xx.x%) | xxx/xxx (xx.x%) | xxx/xxx (xx.x%) |
| Once a day | xxx/xxx (xx.x%) | xxx/xxx (xx.x%) | xxx/xxx (xx.x%) |
| Once every few days | xxx/xxx (xx.x%) | xxx/xxx (xx.x%) | xxx/xxx (xx.x%) |
| Once or twice in the past 2 weeks | xxx/xxx (xx.x%) | xxx/xxx (xx.x%) | xxx/xxx (xx.x%) |
| Not at all in the past 2 weeks | xxx/xxx (xx.x%) | xxx/xxx (xx.x%) | xxx/xxx (xx.x%) |
| Don't know | xxx/xxx (xx.x%) | xxx/xxx (xx.x%) | xxx/xxx (xx.x%) |
| **Vape or e-cigarette at least daily** |  |  |  |
| Yes | xxx/xxx (xx.x%) | xxx/xxx (xx.x%) | xxx/xxx (xx.x%) |
| No | xxx/xxx (xx.x%) | xxx/xxx (xx.x%) | xxx/xxx (xx.x%) |
| **Vaping frequency** |  |  |  |
| Several times a day | xxx/xxx (xx.x%) | xxx/xxx (xx.x%) | xxx/xxx (xx.x%) |
| Once a day | xxx/xxx (xx.x%) | xxx/xxx (xx.x%) | xxx/xxx (xx.x%) |
| Once every few days | xxx/xxx (xx.x%) | xxx/xxx (xx.x%) | xxx/xxx (xx.x%) |
| Once or twice in the past 2 weeks | xxx/xxx (xx.x%) | xxx/xxx (xx.x%) | xxx/xxx (xx.x%) |
| Not at all in the past 2 weeks | xxx/xxx (xx.x%) | xxx/xxx (xx.x%) | xxx/xxx (xx.x%) |
| Don't know | xxx/xxx (xx.x%) | xxx/xxx (xx.x%) | xxx/xxx (xx.x%) |
| **Eats fruit, vegetables daily** |  |  |  |
| Yes | xxx/xxx (xx.x%) | xxx/xxx (xx.x%) | xxx/xxx (xx.x%) |
| No | xxx/xxx (xx.x%) | xxx/xxx (xx.x%) | xxx/xxx (xx.x%) |
| **More than 150 minutes of physical activity per week** |  |  |  |
| Yes | xxx/xxx (xx.x%) | xxx/xxx (xx.x%) | xxx/xxx (xx.x%) |
| No | xxx/xxx (xx.x%) | xxx/xxx (xx.x%) | xxx/xxx (xx.x%) |
| **Knowledge of conditions caused by high blood pressure** |  |  |  |
| Heart attacks | xxx/xxx (xx.x%) | xxx/xxx (xx.x%) | xxx/xxx (xx.x%) |
| Cancer | xxx/xxx (xx.x%) | xxx/xxx (xx.x%) | xxx/xxx (xx.x%) |
| Diabetes | xxx/xxx (xx.x%) | xxx/xxx (xx.x%) | xxx/xxx (xx.x%) |
| High cholesterol | xxx/xxx (xx.x%) | xxx/xxx (xx.x%) | xxx/xxx (xx.x%) |
| Allergy | xxx/xxx (xx.x%) | xxx/xxx (xx.x%) | xxx/xxx (xx.x%) |
| ... |  |  |  |

Note:
Denominators are number of participants with available data.

**10.6 Table 5.**
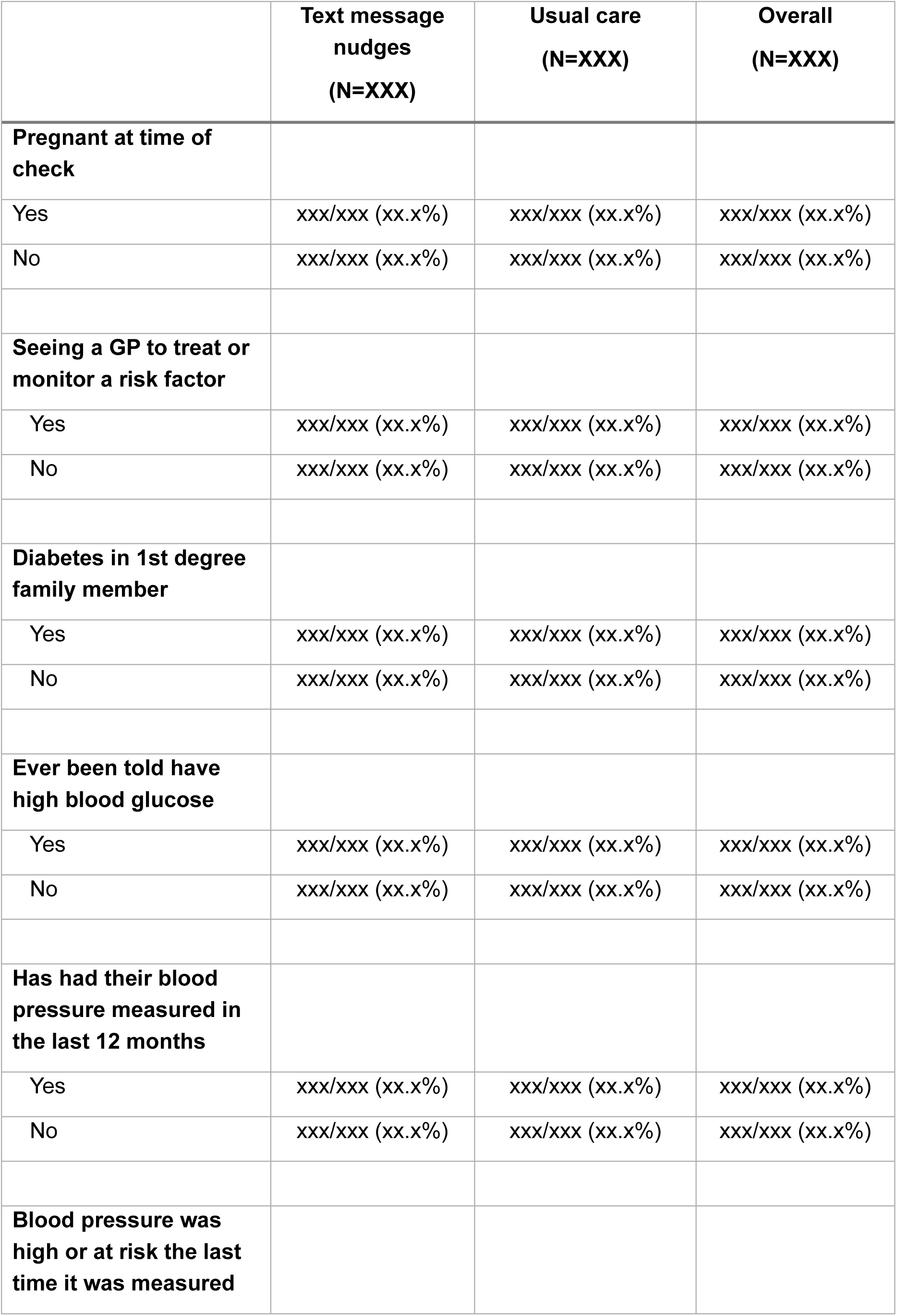
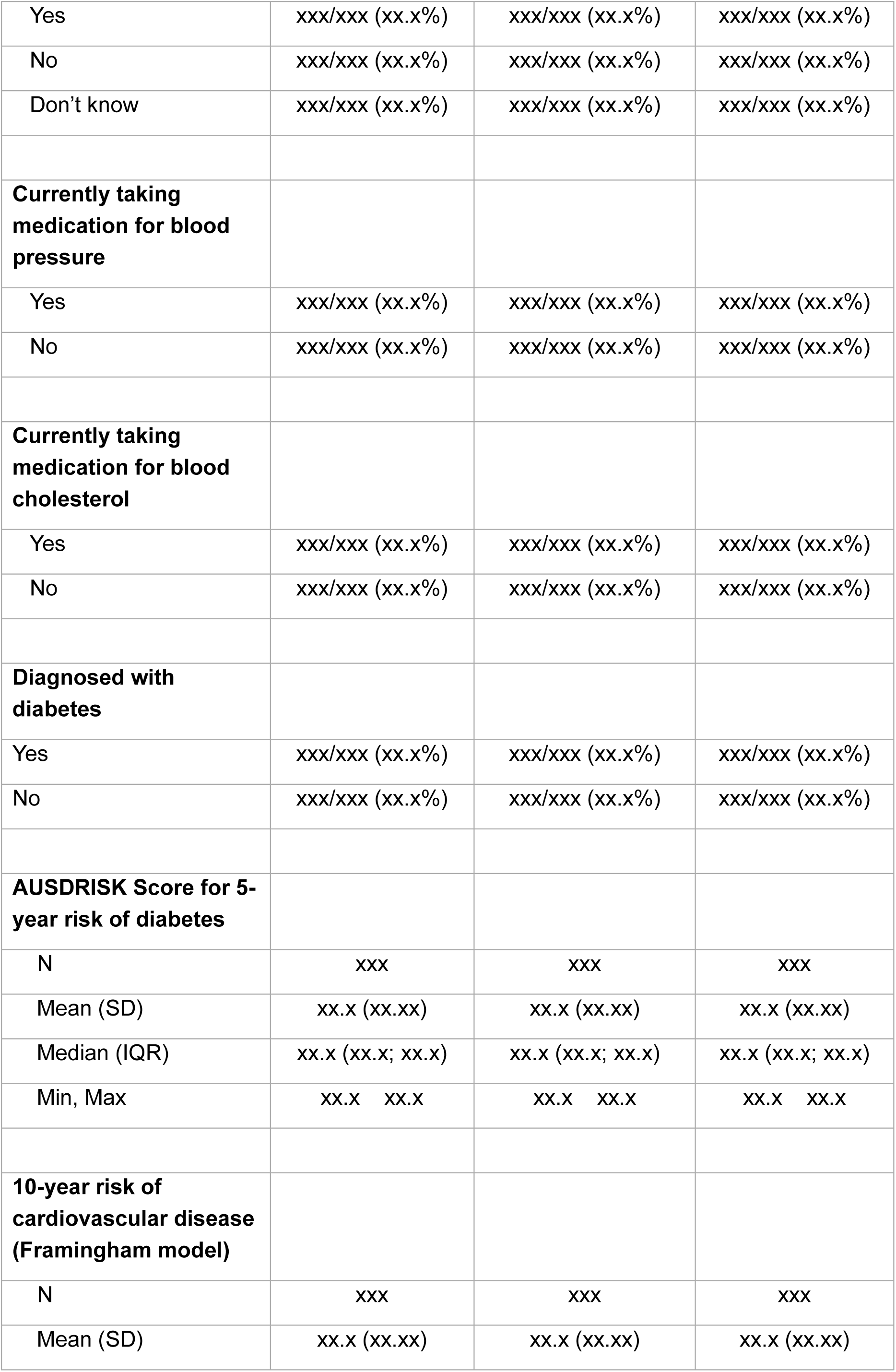
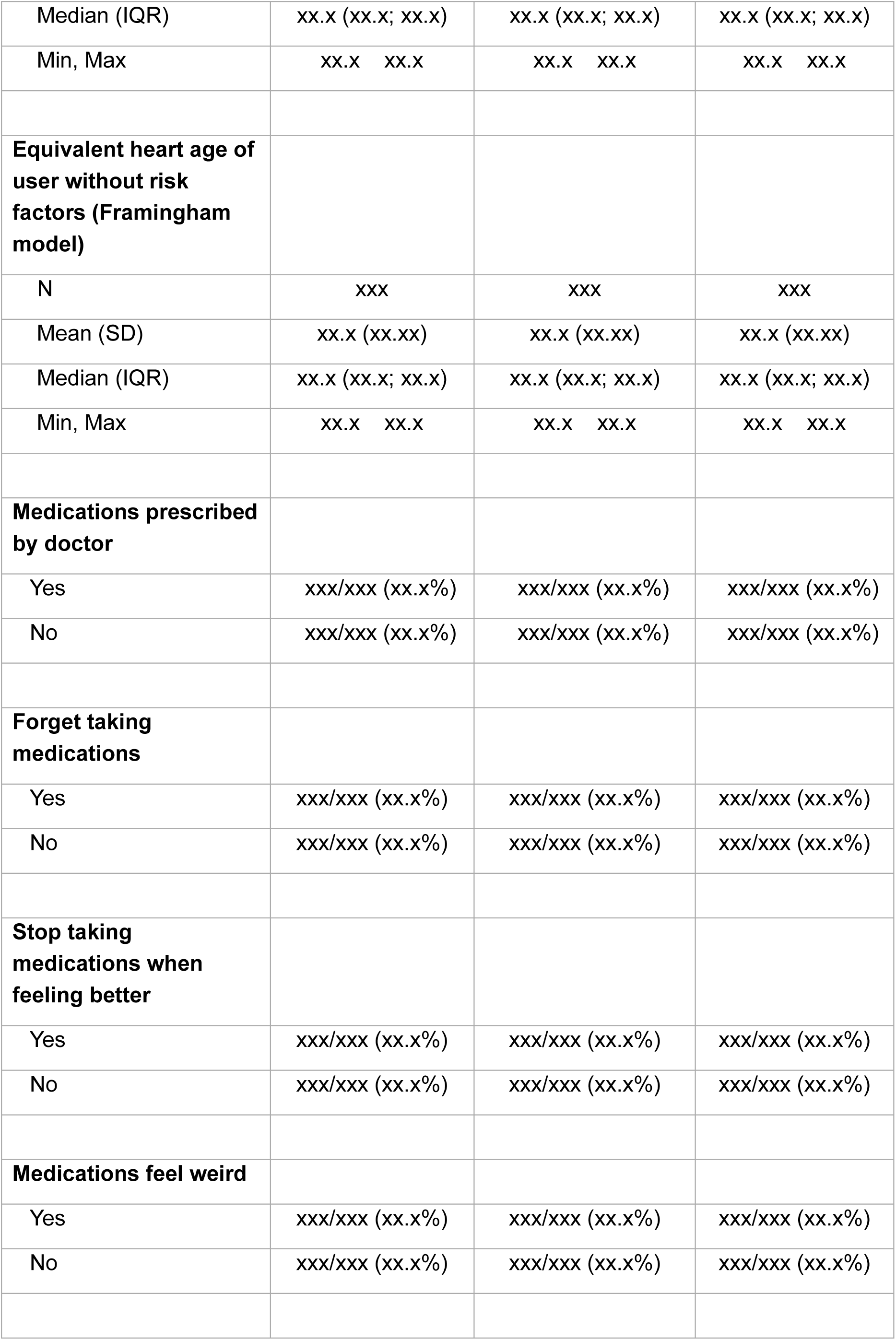
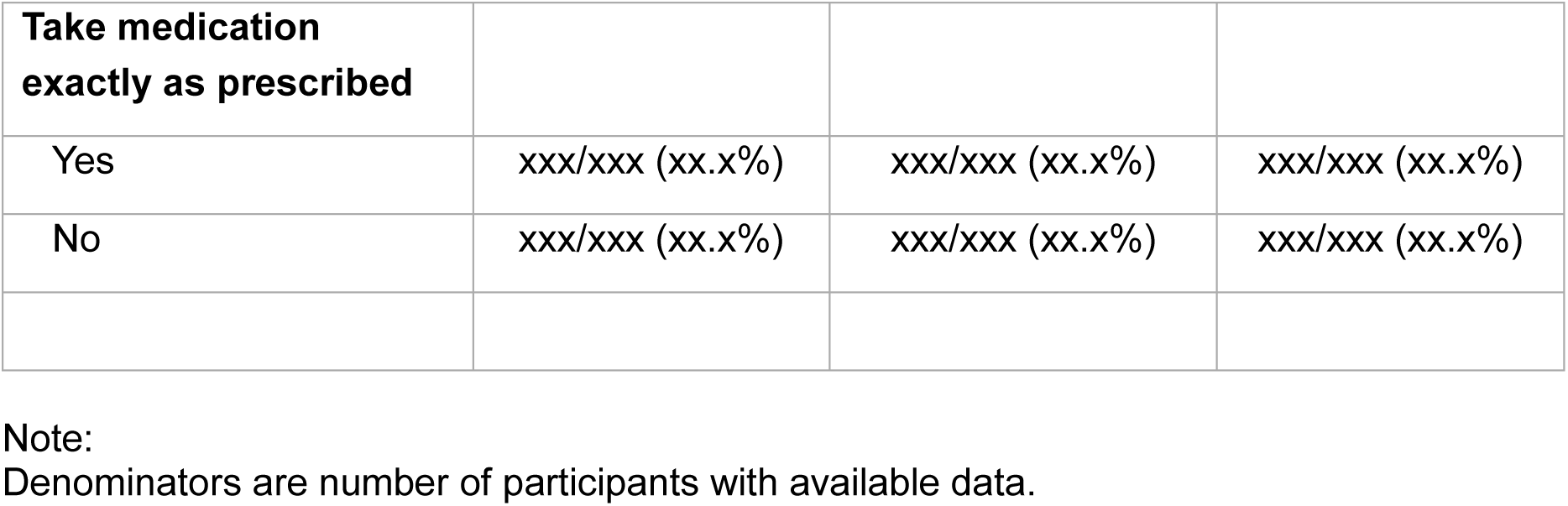
Medical History

**10.7 Table 6.** Physical measurements at baseline

|  | **Text message nudges**<br>**(N=XXX)** | **Usual care**<br>**(N=XXX)** | **Overall**<br>**(N=XXX)** |
| --- | --- | --- | --- |
| **Height (cm)** |  |  |  |
| N | xxx | xxx | xxx |
| Mean (SD) | xx.x (xx.xx) | xx.x (xx.xx) | xx.x (xx.xx) |
| Median (IQR) | xx.x (xx.x; xx.x) | xx.x (xx.x; xx.x) | xx.x (xx.x; xx.x) |
| Min, Max | xx.x xx.x | xx.x xx.x | xx.x xx.x |
| Missing | xxx | xxx | xxx |
| **Weight (kg)** |  |  |  |
| N | xxx | xxx | xxx |
| Mean (SD) | xx.x (xx.xx) | xx.x (xx.xx) | xx.x (xx.xx) |
| Median (IQR) | xx.x (xx.x; xx.x) | xx.x (xx.x; xx.x) | xx.x (xx.x; xx.x) |
| Min, Max | xx.x xx.x | xx.x xx.x | xx.x xx.x |
| Missing | xxx | xxx | xxx |
| **Body Mass Index (kg/m<sup>2</sup>)** |  |  |  |
| N | xxx | xxx | xxx |
| Mean (SD) | xx.x (xx.xx) | xx.x (xx.xx) | xx.x (xx.xx) |
| Median (IQR) | xx.x (xx.x; xx.x) | xx.x (xx.x; xx.x) | xx.x (xx.x; xx.x) |
| Min, Max | xx.x xx.x | xx.x xx.x | xx.x xx.x |
| Missing | xxx | xxx | xxx |
| **Body fat percentage (%)** |  |  |  |
| N | xxx | xxx | xxx |
| Mean (SD) | xx.x (xx.xx) | xx.x (xx.xx) | xx.x (xx.xx) |
| Median (IQR) | xx.x (xx.x; xx.x) | xx.x (xx.x; xx.x) | xx.x (xx.x; xx.x) |
| Min, Max | xx.x xx.x | xx.x xx.x | xx.x xx.x |
| Missing | xxx | xxx | xxx |
| **Diastolic blood pressure (mmHg)** |  |  |  |
| N | xxx | xxx | xxx |
| Mean (SD) | xx.x (xx.xx) | xx.x (xx.xx) | xx.x (xx.xx) |
| Median (IQR) | xx.x (xx.x; xx.x) | xx.x (xx.x; xx.x) | xx.x (xx.x; xx.x) |
| Min, Max | xx.x xx.x | xx.x xx.x | xx.x xx.x |
| Missing | xxx | xxx | xxx |
| **Systolic blood pressure (mmHg)** |  |  |  |
| N | xxx | xxx | xxx |
| Mean (SD) | xx.x (xx.xx) | xx.x (xx.xx) | xx.x (xx.xx) |
| Median (IQR) | xx.x (xx.x; xx.x) | xx.x (xx.x; xx.x) | xx.x (xx.x; xx.x) |
| Min, Max | xx.x xx.x | xx.x xx.x | xx.x xx.x |
| Missing | xxx | xxx | xxx |
| **Heart rate** |  |  |  |
| N | xxx | xxx | xxx |
| Mean (SD) | xx.x (xx.xx) | xx.x (xx.xx) | xx.x (xx.xx) |
| Median (IQR) | xx.x (xx.x; xx.x) | xx.x (xx.x; xx.x) | xx.x (xx.x; xx.x) |
| Min, Max | xx.x xx.x | xx.x xx.x | xx.x xx.x |
| Missing | xxx | xxx | xxx |

**10.8 Figure 2.**
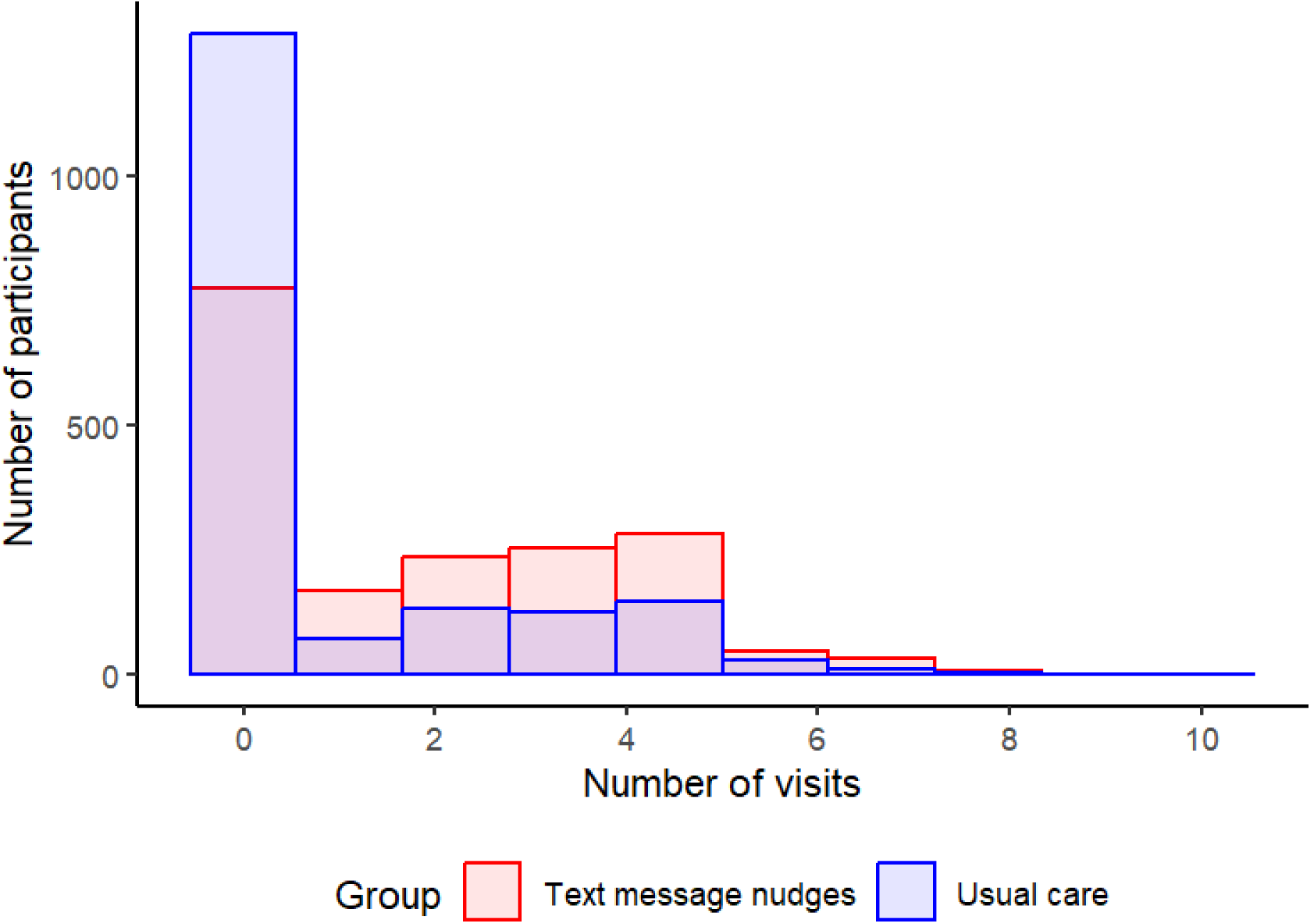
Number of checks per user (illustration)

**10.9 Table 7.**
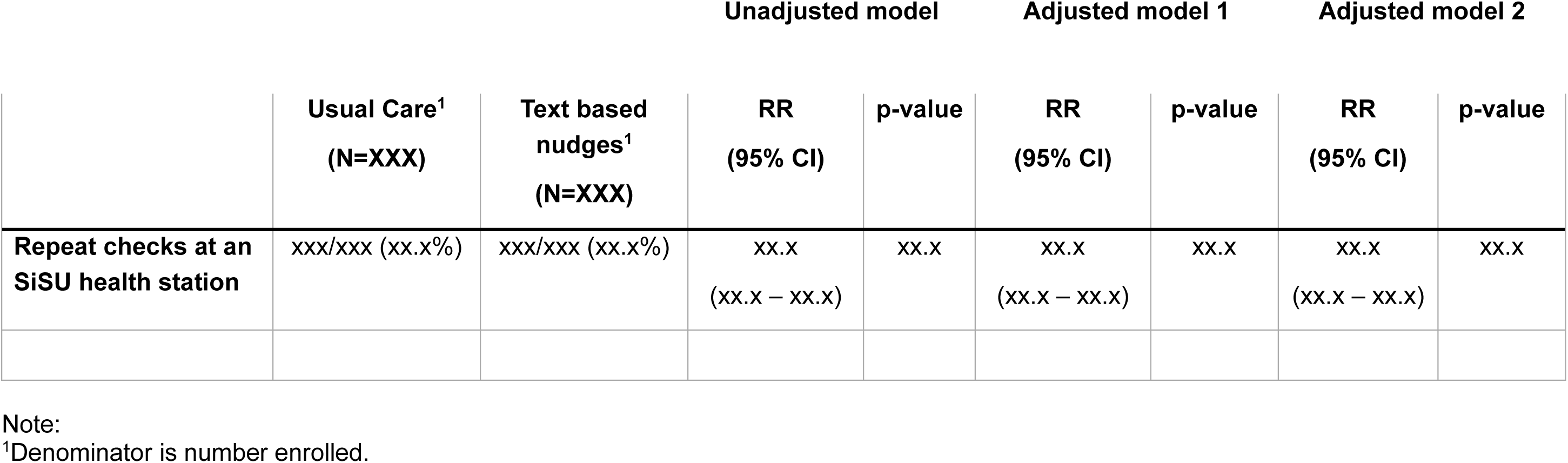
Primary outcome

|  |  |  | Unadjusted model |  | Adjusted model 1 |  | Adjusted model 2 |  |
| --- | --- | --- | --- | --- | --- | --- | --- | --- |
|  | Usual Care<sup>1</sup><br>(N=XXX) | Text based nudges<sup>1</sup><br>(N=XXX) | RR<br>(95% CI) | p-value | RR<br>(95% CI) | p-value | RR<br>(95% CI) | p-value |
| Repeat checks at an SiSU health station | xxx/xxx (xx.x%) | xxx/xxx (xx.x%) | xx.x<br>(xx.x – xx.x) | xx.x | xx.x<br>(xx.x – xx.x) | xx.x | xx.x<br>(xx.x – xx.x) | xx.x |

Note: <sup>1</sup>Denominator is number enrolled.

**10.10 Table 8.** Binary secondary outcomes

|  | **Text message nudges<sup>1</sup>**<br>**(N=XXX)** | **Usual care<sup>1</sup>**<br>**(N=XXX)** | **RR**<br>**(95% CI)** | **p-value** |
| --- | --- | --- | --- | --- |
| **Achieve target BP** | xxx/xxx (xx.x%) | xxx/xxx (xx.x%) | xx.x<br>(xx.x – xx.x) | xx.x |
| **Change in use of BP medications** | xxx/xxx (xx.x%) | xxx/xxx (xx.x%) | xx.x<br>(xx.x – xx.x) | xx.x |
| **Change in health risk behaviours** | xxx/xxx (xx.x%) | xxx/xxx (xx.x%) | xx.x<br>(xx.x – xx.x) | xx.x |
| **Self-reported medical attendance with GP** | xxx/xxx (xx.x%) | xxx/xxx (xx.x%) | xx.x<br>(xx.x – xx.x) | xx.x |
| <each level of variable> | xxx/xxx (xx.x%) | xxx/xxx (xx.x%) | xx.x<br>(xx.x – xx.x) | xx.x |
| **Awareness of high BP risks** | xxx/xxx (xx.x%) | xxx/xxx (xx.x%) | xx.x<br>(xx.x – xx.x) | xx.x |

Note:
<sup>1</sup>Denominator is number enrolled.

**10.11 Table 9.**
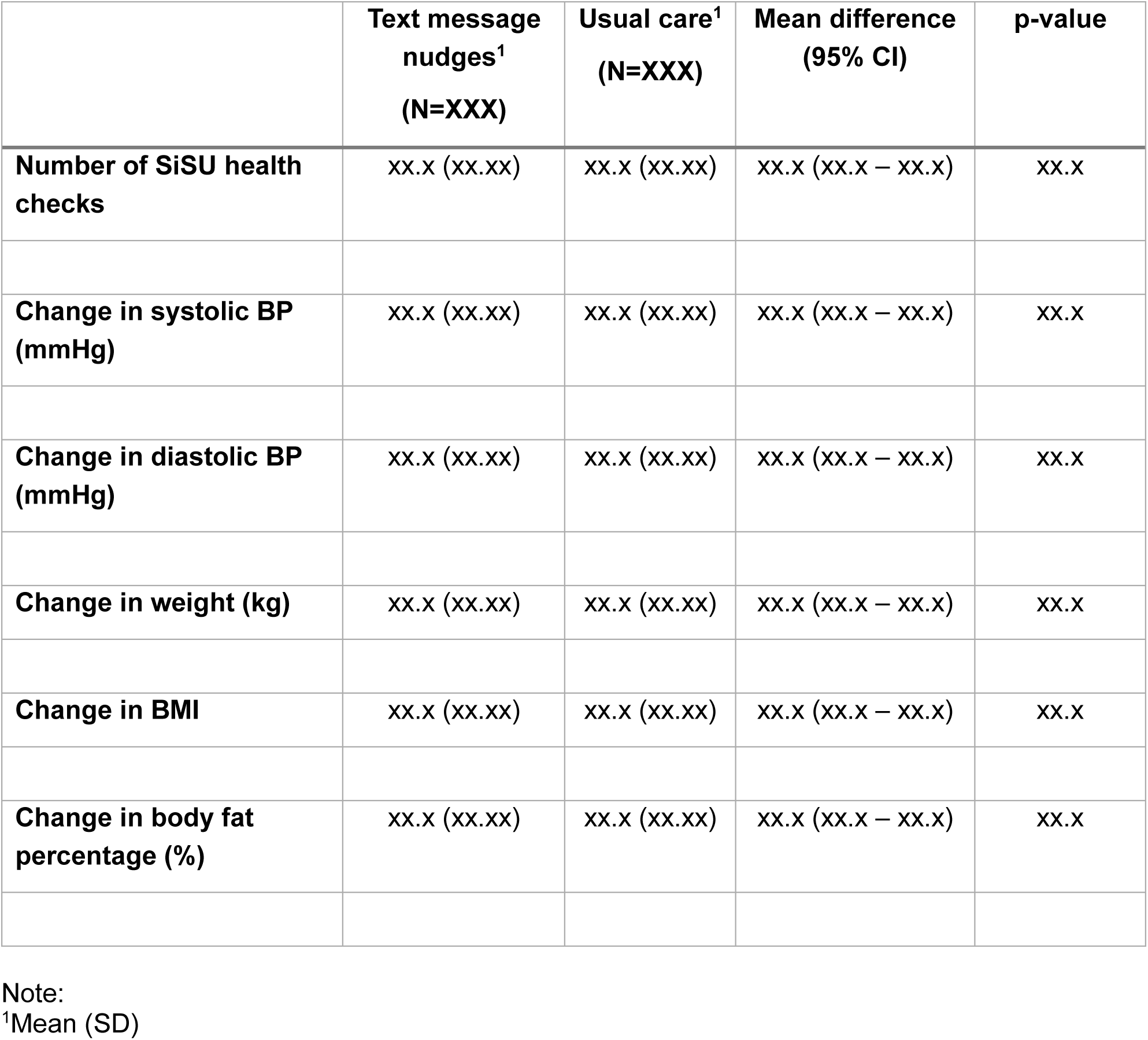
Continuous secondary outcomes

|  | **Text message nudges<sup>1</sup>**<br>**(N=XXX)** | **Usual care<sup>1</sup>**<br>**(N=XXX)** | **Mean difference**<br>**(95% CI)** | **p-value** |
| --- | --- | --- | --- | --- |
| **Number of SiSU health checks** | xx.x (xx.xx) | xx.x (xx.xx) | xx.x (xx.x – xx.x) | xx.x |
| **Change in systolic BP (mmHg)** | xx.x (xx.xx) | xx.x (xx.xx) | xx.x (xx.x – xx.x) | xx.x |
| **Change in diastolic BP (mmHg)** | xx.x (xx.xx) | xx.x (xx.xx) | xx.x (xx.x – xx.x) | xx.x |
| **Change in weight (kg)** | xx.x (xx.xx) | xx.x (xx.xx) | xx.x (xx.x – xx.x) | xx.x |
| **Change in BMI** | xx.x (xx.xx) | xx.x (xx.xx) | xx.x (xx.x – xx.x) | xx.x |
| **Change in body fat percentage (%)** | xx.x (xx.xx) | xx.x (xx.xx) | xx.x (xx.x – xx.x) | xx.x |

Note:
<sup>1</sup>Mean (SD)

**10.12 Figure 3.**
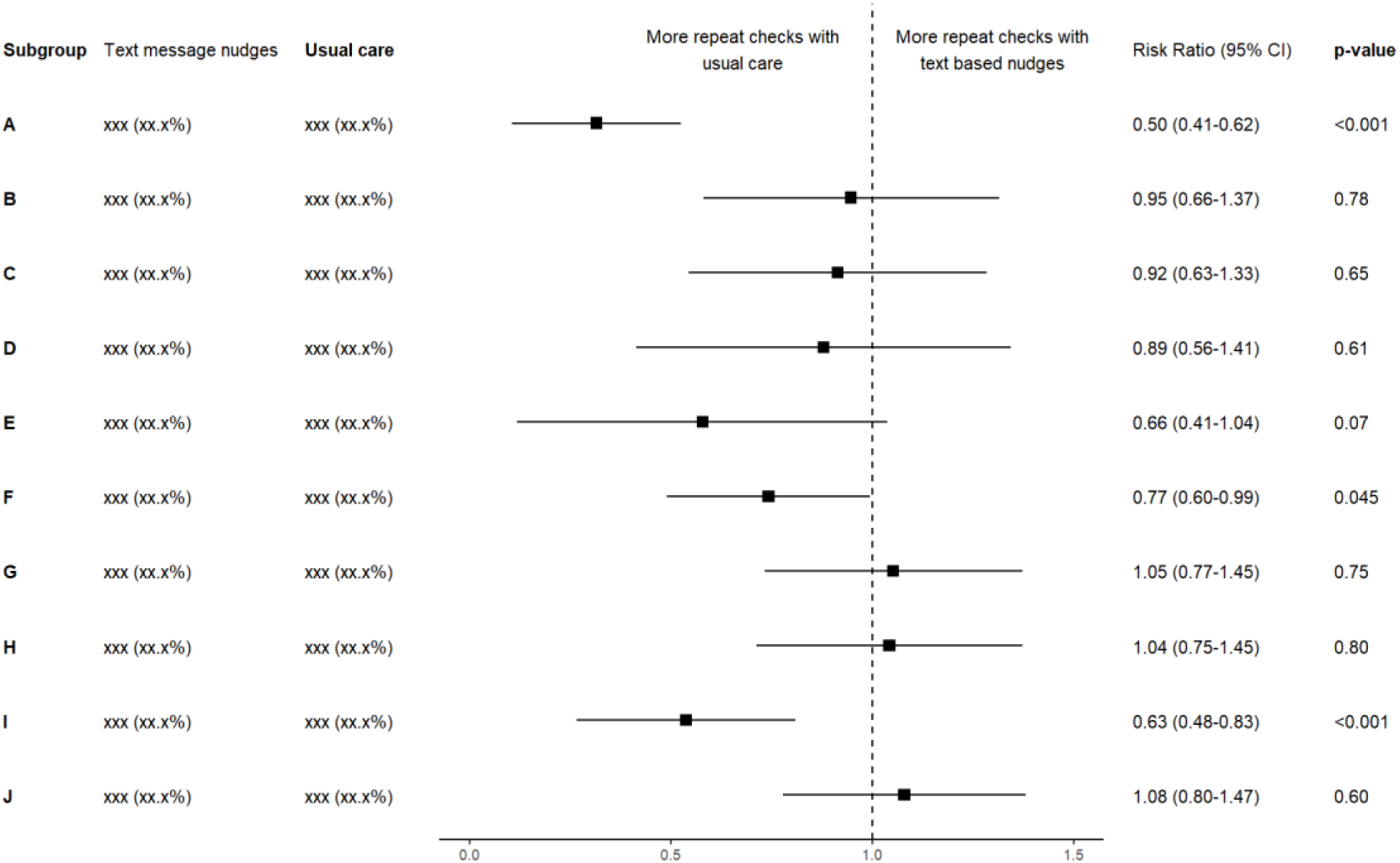
Subgroup analyses (illustration)

